# Concomitant Use of VA-ECMO and Impella Support for Cardiogenic Shock

**DOI:** 10.1101/2023.07.24.23293127

**Authors:** Shan P. Modi, Yeahwa Hong, McKenzie M. Sicke, Nicholas R. Hess, Wyatt J. Klass, Luke A. Ziegler, Ryan M. Rivosecchi, Gavin W. Hickey, David J. Kaczorowski, Raj Ramanan

## Abstract

**Background:** VA-ECMO with concomitant Impella support (ECpella) is an emerging treatment modality for cardiogenic shock (CS). Survival outcomes by CS etiology with ECpella support have not been well-described.

**Methods:** This study was a retrospective, single-center analysis of patients with cardiogenic shock due to acute myocardial infarction (AMI-CS) or decompensated heart failure (ADHF-CS) supported with ECpella from December 2020 to January 2023. Primary outcomes included 90-day survival post-discharge and destination after support. Secondary outcomes included complications post-ECpella support.

**Results:** A total of 44 patients were included (AMI-CS, *n =* 20, and ADHF-CS, *n* = 24). Patients with AMI-CS and ADHF-CS had similar survival 90 days post-discharge (*p*= .267) with similar destinations after ECpella support (*p =* .220). Limb ischemia and acute kidney injury occurred more frequently in patients presenting with AMI-CS (*p=.*013; *p* = .030). Patients with initial Impella support were more likely to survive ECpella support and be bridged to transplant (*p*=.033) and less likely to have a cerebrovascular accident *(p*=.016). Sub-analysis of ADHF-CS patients into acute-on-chronic decompensated heart failure and de novo heart failure demonstrated no difference in survival or destination.

**Conclusion:** ECpella can be used to successfully manage patients with CS. There is no difference in survival or destination for AMI-CS and ADHF-CS in patients with ECpella support. Patients with initial Impella support are more likely to survive ECpella support and bridge to transplant. Future multicenter studies are required to fully analyze the differences between AMI-CS and ADHF-CS with ECpella support.

**Clinical Perspectives:** **What is New?**

ECpella support is a feasible support strategy for allcomers in severe cardiogenic shock. This study demonstrates that ECpella can be utilized not only as a salvage therapy and venting strategy for those in cardiogenic shock on VA-ECMO, but also can be utilized as a method for additional cardiac support for patients with initial Impella support. There were no differences in survival between cardiogenic shock secondary to acute myocardial infarction and cardiogenic secondary to acute decompensated heart failure.

**What are the clinical implications?**

Although ECpella patients that received initial Impella support have higher success in bridging to heart transplant, allcomers on ECpella support should be evaluated for advanced therapies early in their clinical course. Further studies are required to ascertain the differences in pathophysiology between cardiogenic shock secondary to acute myocardial infarction and cardiogenic secondary to acute decompensated heart failure and determine appropriate support strategies for differing cardiogenic shock phenotypes.

## Introduction

Cardiogenic shock (CS) is a complex syndrome associated with low cardiac output secondary to dysfunctional myocardium leading to end-organ hypoperfusion, systemic vasoconstriction, and generalized hypoxia. Early recognition with initiation of inotropic and/or mechanical support is essential in the management of CS^1^. However, the outcomes of patients with CS remain poor with short-term mortality exceeding 50%, despite early coronary revascularization in the setting of acute myocardial infarction^1,2^. Over the past decade, temporary mechanical circulatory support devices have emerged as a pivotal component of CS management^3^. Among the available technologies, both venoarterial-extracorporeal membrane oxygenation (VA-ECMO) and Impella (Abiomed, Danvers, MA) are increasingly employed in patients with severe CS^2,4^.

The use of VA-ECMO has grown exponentially over the past decade in the United States, especially in the setting of CS secondary to AMI^5^. Peripheral VA-ECMO is able to provide full circulatory and respiratory support, although the retrograde nature of the blood flow towards the heart leads to increased left ventricular afterload and wall stress^6^. In the setting of severely impaired left ventricular contractility, the left ventricle may become pressurized with increased end-diastolic volume causing increased myocardial oxygen demand^7^. Furthermore, overall coronary perfusion may become compromised due to high diastolic ventricular pressures^8^. The incidence of left ventricular overload from VA-ECMO varies widely, but the previously reported rate is as high as 70%^8,9^. With emerging evidence suggesting adverse consequences of left ventricular overload, the topic of left ventricular unloading strategies is an area of active investigation^9^.

Of the various available strategies, the concomitant use of a microaxial left ventricular assist device (Impella) to unload the left ventricle in patients supported with VA-ECMO (ECpella) has been increasingly utilized. Prior studies evaluating the impact of ECpella in patients with CS have collectively demonstrated favorable outcomes^10,11^. Many of these prior studies were limited to partial flow, femorally inserted Impella devices, such as the Impella 2.5 and CP, with shorter support duration^12^. Therefore, the impact of larger Impella devices, such as the Impella 5.5, for longer support duration is rather limited. In this study, we aim to evaluate CS patients with ECpella support with Impella CP or Impella 5.5 and stratify outcomes by both cardiogenic shock etiology and initial support strategy.

## Materials and Methods

This single center, retrospective cohort study was approved by the local institutional review board (IRB #18120143) at the University of Pittsburgh Medical Center, Pittsburgh, PA and performed in accordance with the principles set forth by the Declaration of Helsinki. This study was completed at the University of Pittsburgh Presbyterian Hospital, an academic tertiary center in the University of Pittsburgh Medical Center (UPMC) Health System. The need for informed consent was waived by our local institutional review board due to the retrospective nature of the study.

### Study Population

Patients were identified through internal review of an internal ECMO and Impella database maintained by perfusion services at University of Pittsburgh Presbyterian Hospital between December 2020 to January 2023. Adult patients (18 years or older) that received simultaneous support with an Impella CP or Impella 5.5 (Abiomed; Danvers, MA) axial flow pump and veno-arterial ECMO were included in the study. Determination of concomitant support for patients declining from CS was made collectively by adjudication via a diverse, multidisciplinary physician team of a cardiothoracic intensivist, cardiothoracic surgeon, and an advanced heart failure cardiologist. Patients requiring simultaneous support for post-cardiotomy shock were excluded from the study. Systemic anticoagulation with bivalirudin was provided to all patients in concordance with protocol at the study center. All patients received at least 24 hours of simultaneous Impella and ECMO support to be included in the analysis.

### Study Variables

All baseline demographic characteristics and laboratory variables related to mechanical circulatory support were obtained via the internal ECMO and Impella database and cross-referenced with manual review of the electronic medical record. Patients with ECpella support were classified by CS etiology including CS secondary to acute myocardial infarction (AMI-CS) or cardiogenic shock secondary to acute decompensated heart failure (ADHF-CS). Society of Cardiovascular Angiography and Intervention (SCAI) stages are reported from initial patient presentation using the defined criteria by the SCAI consensus statement^13^.

### Clinical Endpoints

The primary outcomes assessed in this study were 90-day survival and destination after ECpella support stratified by CS etiology. Destinations included death, bridge to recovery, bridge to LVAD, and bridge to heart transplant. Secondary outcomes included length of stay (LOS) and complications during ECpella support including major and minor bleeding per TIMI criteria, limb ischemia requiring surgical intervention, deep vein thrombosis (DVT) or pulmonary embolism (PE), ischemic and hemorrhagic cerebrovascular accidents (CVA), infection, acute kidney injury (AKI), necessity for continuous renal replacement therapy (CRRT) or continuous veno-venous hemodialysis (CVVHD), heparin-induced thrombocytopenia (HIT) and Impella pump thrombosis. Primary and secondary outcomes were also assessed in sub-analysis groups including patient stratification based on initial support strategy and by acute heart failure etiology (de novo heart failure versus acute-on-chronic decompensated heart failure).

### Statistical Analysis

Data is presented as frequency (percentage) for categorical variables, mean (± standard deviation) for Gaussian continuous variables and median [IQR] for non-Gaussian continuous variables. Pearson’s Chi-square test was utilized for categorical comparisons with Fisher’s exact test utilized for group sizes with a n ≤ 5. Post-hoc analysis via Pearson adjusted residuals were utilized to delineate associations in significant categorical comparisons. Student’s *t*-test was employed for parametric continuous variables with Wilcoxon-Rank Sum (Mann-Whitney ***U***) test employed for non-parametric variables. Shapiro-Wilk’s test was applied to all continuous variables to assess for normality. Kaplan-Meier survival estimate curves were calculated with freedom from mortality and assessed by two-sided log-rank test. Analyses were performed via STATA SE (Version 17.0, College Station, TX). Statistical significance was defined as two-sided *p* < 0.05 for all tests.

## Results

### Baseline Characteristics

45 patients with CS treated with ECpella support were identified in our registry with 1 patient with post-cardiotomy shock excluded from analysis. 20 patients (45.5%) sustained CS secondary to AMI-CS. Baseline characteristics are summarized in **Table 1**. There was no significant difference between the age of the patients in each group and the majority of the patients analyzed were white males. No difference was ascertained in the socioeconomic status of each cohort as determined by the distressed communities index (DCI). The majority of patients in our study were initially transferred from an outside hospital with similar SCAI stages on initial presentation. Patients that presented with AMI-CS were more likely to be diabetic (AMI-CS: 70.0%, ADHF-CS: 25.0%, *P =* .003) and have a prior history of coronary artery disease (AMI-CS: 85.0%; ADHF-CS: 33.3, *P* = .001). In addition, patients that presented with AMI-CS were more likely to have coronary intervention (percutaneous coronary intervention or coronary artery bypass) during their admission (AMI-CS: 90.0%, ADHF-CS: 12.5%, *P* <.001).

**Table 1.** Baseline Characteristics by CS Etiology.

| Patient Demographics |  |  |  |  |
| --- | --- | --- | --- | --- |
| Variable | All (n=44) | AMI-CS (n=20) | ADHF-CS (n=24) | P |
| Male | 27 (61.4) | 13 (65.0) | 14 (58.3) | .651 |
| Age (y) | 51.8 ± 13.0 | 54.5 ± 9.9 | 49.6 ± 14.9 | .224 |
| Race |  |  |  | .160 |
| White | 35 (79.5) | 15 (75.0) | 20 (83.3) |  |
| Black | 2 (4.5) | 0 (0) | 2 (8.3) |  |
| Not Specified | 7 (16.0) | 5 (25.0) | 2 (8.3) |  |
| DCI | 44.2 ± 26.7 | 49.2 ± 27.1 | 40.0 ± 26.3 | .264 |
| Transfer | 36 (81.8) | 17 (85.0) | 19 (79.2) | .617 |
| SCAI Stage Prior to MCS |  |  |  | .557 |
| C | 8 (18.2) | 4 (20.0) | 4 (16.7) |  |
| D | 17 (38.6) | 6 (30.0) | 11 (45.8) |  |
| E | 19 (43.2) | 10 (50.0) | 9 (37.4) |  |
| Comorbidities |  |  |  |  |
| HTN | 23 (52.3) | 12 (60.0) | 11 (45.8) | .349 |
| DM | 20 (45.5) | 14 (70.0) | 6 (25.0) | .003 |
| CKD | 8 (18.2) | 2 (10.0) | 6 (25.0) | .199 |
| CAD | 25 (56.8) | 17 (85.0) | 8 (33.3) | .001 |
| CVA/TIA | 2 (4.6) | 1 (5.0) | 1 (4.2) | .895 |
| DVT/PE | 1 (2.3) | 0 (0.0) | 1 (4.2) | .356 |
| Atrial Fibrillation | 3 (6.8) | 1 (5.0) | 2 (8.3) | .662 |
| PAD | 2 (4.6) | 0 (0.0) | 2 (8.3) | .186 |
| COPD | 3 (6.8) | 1 (5.0) | 2 (8.3) | .662 |
| Coronary Intervention and Mechanical Support |  |  |  |  |
| Impella Type |  |  |  | .908 |
| Impella CP | 15 (34.1) | 7 (35.0) | 8 (33.3) |  |
| Impella 5.5 | 29 (65.9) | 13 (65.0) | 16 (66.7) |  |
| Days to Impella Placement | 1 [0-3] | 1[0-2.5] | 1.5 [0-5.5] | .177 |
| Days of Impella Support | 11[6-22.5] | 11.5 [7-29.5] | 11 [6-15.5] | .555 |
| Initial VA-ECMO Support | 21 (47.7) | 11 (55) | 10 (41.7) | .378 |
| Coronary Intervention | 21 (47.7) | 18 (90.0) | 3 (12.5) | <.001 |
| Labs Prior To ECpella Support |  |  |  |  |
| Lowest pH | 7.30 [7.22-7.36] | 7.30 [7.20-7.34] | 7.33 [7.22-7.40] | .317 |
| Highest Lactate (mmol/L) | 4.3 [2.1-9.6] | 9.1 [2.75-11.5] | 3.3 [1.9-7.1] | .113 |
| Lowest Hemoglobin (g/dL) | 10.6 ± 2.9 | 10.2 ± 3.1 | 11.0 ± 2.8 | .380 |
| Lowest Platelets (10<sup>9</sup>/L) | 151 [99-237] | 166.5 [95-261] | 136 [102-216] | .671 |
| **Highest Creatinine (mg/dL)** | 1.5 [1.1-2.4] | 1.6 [1.2-2.7] | 1.4 [1.1-2.4] | .629 |
| **Highest Total Bilirubin (mg/dL)** | 1.4 [.6-2.4] | 1.1 [.7-1.5] | 1.4 [.6-2.6] | .398 |

CAD = Coronary Artery Disease, CKD= Chronic Kidney Disease, COPD=Chronic Obstructive Pulmonary Disease, CVA=Cerebrovascular Accident, DCI= Distressed Communities Index, DVT=Deep Vein Thrombosis, PAD=Peripheral Artery Disease, PE=Pulmonary Embolism, SCAI= Society of Cardiovascular Angiography and Intervention, TIA=Transient Ischemic Attack

Baseline characteristics stratified by initial support strategy are summarized in **Table 2**. There were no significant differences in age, gender, race, socioeconomic status, transfer status, or SCAI stage prior to mechanical circulatory support (MCS) between both cohorts. Patients that had initial Impella support were more likely to have chronic kidney disease compared to those with initial VA-ECMO support (Impella: 30.4%, VA-ECMO: 4.8%, *P =* .027). Patients with initial VA-ECMO support were more likely to have ECpella support with Impella 5.5 compared to patients with initial Impella support (Impella: 52.2%, VA-ECMO: 81.0%, *P =* .044). Prior to full ECpella support, patients with initial VA-ECMO support had lower hemoglobin counts (Impella: 11.8 [10.0-13.3], VA-ECMO: 9.0 [7.5-10.5], *P* = .021) and lower platelet counts (Impella: 203 [128-268], VA-ECMO: 97 [84-166], *P* = .002) compared to initial Impella support.

**Table 2.** Baseline Characteristics by Initial Support Strategy.

| **Patient Demographics** |  |  |  |
| --- | --- | --- | --- |
| **Variable** | **Initial Impella Support**<br>**(n=23)** | **Initial ECMO Support**<br>**(n=21)** | **P** |
| **CS Etiology** |  |  | .378 |
| AMI | 9 (39.1) | 11 (52.4) |  |
| ADHF | 14 (60.9) | 10 (47.6) |  |
| **Age (y)** | 52.5 ± 13.6 | 51.1 ± 12.6 | .723 |
| **Race** |  |  | .857 |
| White | 19 (82.6) | 16 (76.2) |  |
| Black | 1 (4.3) | 1 (4.8) |  |
| Not Specified | 3 (13.0) | 4 (19.0) |  |
| **DCI** | 42.7 ± 27.1 | 45.8 ± 26.9 | .711 |
| **Transfer** | 17 (73.9) | 19 (90.5) | .155 |
| **SCAI Stage Prior to MCS** |  |  | .202 |
| C | 5 (21.7) | 3 (14.3) |  |
| D | 11 (47.8) | 6 (28.6) |  |
| E | 7 (30.4) | 12 (57.1) |  |
| **Comorbidities** |  |  |  |
| **HTN** | 11 (47.8) | 12 | .537 |
| **DM** | 8 (34.8) | 12 | .137 |
| **CKD** | 7 (30.4) | 1 (4.8) | **.027** |
| **CAD** | 13 | 12 | .967 |
| **CVA/TIA** | 1 | 1 | .947 |
| **DVT/PE** | 1 | 0 | .334 |
| **Atrial Fibrillation** | 2 | 1 | .605 |
| **PAD** | 2 | 0 | .167 |
| **COPD** | 1 | 2 | .496 |
| **Coronary Intervention and Mechanical Support** |  |  |  |
| **Impella Type** |  |  | **.044** |
| Impella CP | 11 (47.8) | 4 (19.0) |  |
| Impella 5.5 | 12 (52.2) | 17 (81.0) |  |
| **Days to Impella Placement** | 0 [0-3] | 1 [1-3] | .140 |
| **Impella Support Duration** | 13 [6-29] | 10 [6-16] | .689 |
| **Coronary Intervention** | 9 (39.1) | 12 (57.1) | .232 |
| **Labs Prior To ECPella Support** |  |  |  |
| **Lowest pH** | 7.30 [7.22-7.34] | 7.3 [7.22-7.38] | .952 |
| **Highest Lactate (mmol/L)** | 3.7 [2.3-8.0] | 6.7 [1.9-11.5] | .661 |
| **Lowest Hemoglobin (g/dL)** | 11.8 [10.0-13.3] | 9 [7.5-10.5] | **.021** |
| **Lowest Platelets (10<sup>9</sup>/L)** | 203 [128-268] | 97 [84-166] | **.002** |
| **Highest Creatinine (mg/dL)** | 1.4 [1.1-2.2] | 1.8 [1.0-2.5] | .842 |
| **Highest Total Bilirubin (mg/dL)** | 1.4 [.6-2.5] | 1.3 [.7-2.0] | .968 |

In a sub analysis stratifying ADHF-CS patients into cohorts with acute-on-chronic decompensated heart failure (ACDHF) and de novo heart failure, 12 patients (50.0%) presented with de novo acute heart failure (**Table 3**). Patients with ACDHF were more likely to be older (ACHDF: 57.8 ± 10.4, de novo: 41.4 ± 14.6; *P* = .044) and male (ACHDF: 91.7%, de novo: 25.0%, *P* = .001). There were no significant differences in initial support strategy, Impella type, time to Impella placement, or Impella support duration. Patients with ACDHF were more likely to have a history of hypertension (ACDHF: 66.7%, de novo: 25.0%, *P* = .041) and chronic kidney disease (ACDHF: 50.0%, de novo: 0.0%, *P* = .005). No significant differences were ascertained in baseline laboratory values prior to ECpella support between both cohorts.

**Table 3.**
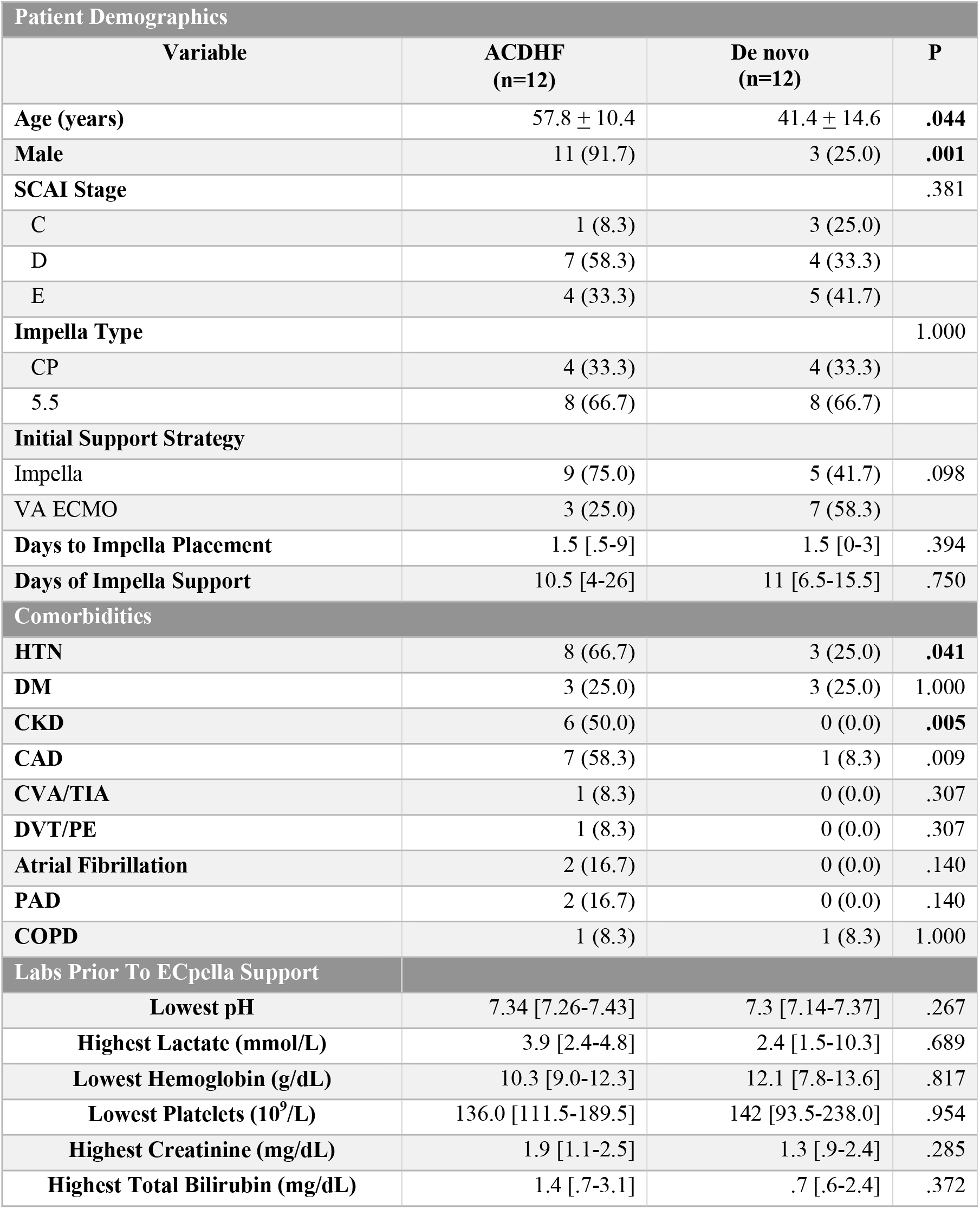
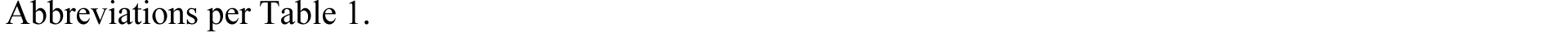
Baseline Characteristics: ACDHF vs. de novo HF.

| Patient Demographics |  |  |  |
| --- | --- | --- | --- |
| Variable | ACDHF<br>(n=12) | De novo<br>(n=12) | P |
| **Age (years)** | 57.8 ± 10.4 | 41.4 ± 14.6 | **.044** |
| **Male** | 11 (91.7) | 3 (25.0) | **.001** |
| **SCAI Stage** |  |  | .381 |
| C | 1 (8.3) | 3 (25.0) |  |
| D | 7 (58.3) | 4 (33.3) |  |
| E | 4 (33.3) | 5 (41.7) |  |
| **Impella Type** |  |  | 1.000 |
| CP | 4 (33.3) | 4 (33.3) |  |
| 5.5 | 8 (66.7) | 8 (66.7) |  |
| **Initial Support Strategy** |  |  |  |
| Impella | 9 (75.0) | 5 (41.7) | .098 |
| VA ECMO | 3 (25.0) | 7 (58.3) |  |
| **Days to Impella Placement** | 1.5 [.5-9] | 1.5 [0-3] | .394 |
| **Days of Impella Support** | 10.5 [4-26] | 11 [6.5-15.5] | .750 |
| Comorbidities |  |  |  |
| **HTN** | 8 (66.7) | 3 (25.0) | **.041** |
| **DM** | 3 (25.0) | 3 (25.0) | 1.000 |
| **CKD** | 6 (50.0) | 0 (0.0) | **.005** |
| **CAD** | 7 (58.3) | 1 (8.3) | .009 |
| **CVA/TIA** | 1 (8.3) | 0 (0.0) | .307 |
| **DVT/PE** | 1 (8.3) | 0 (0.0) | .307 |
| **Atrial Fibrillation** | 2 (16.7) | 0 (0.0) | .140 |
| **PAD** | 2 (16.7) | 0 (0.0) | .140 |
| **COPD** | 1 (8.3) | 1 (8.3) | 1.000 |
| Labs Prior To ECPella Support |  |  |  |
| **Lowest pH** | 7.34 [7.26-7.43] | 7.3 [7.14-7.37] | .267 |
| **Highest Lactate (mmol/L)** | 3.9 [2.4-4.8] | 2.4 [1.5-10.3] | .689 |
| **Lowest Hemoglobin (g/dL)** | 10.3 [9.0-12.3] | 12.1 [7.8-13.6] | .817 |
| **Lowest Platelets (10<sup>9</sup>/L)** | 136.0 [111.5-189.5] | 142 [93.5-238.0] | .954 |
| **Highest Creatinine (mg/dL)** | 1.9 [1.1-2.5] | 1.3 [.9-2.4] | .285 |
| **Highest Total Bilirubin (mg/dL)** | 1.4 [.7-3.1] | .7 [.6-2.4] | .372 |

Abbreviations per Table 1.

### Clinical Endpoints

The overall 90-day survival for all patients with ECpella support was 54.6% **(Table 4)**. There was no statistically significant difference in 90-day post-discharge survival among patients with AMI-CS and ADHF-CS (AMI-CS: 45.0%, ADHF-CS: 62.5%, *P* = .267). Stratification by initial support strategy also demonstrated no statistically significant difference in 90-day survival **(Table 5)** between Impella and VA ECMO groups (Impella: 65.2%, VA-ECMO: 42.9%, *P* = .120). In the ADHF-CS cohort, comparison between patients with ACDHF and de novo heart failure **(Table 6)** did not detect any significant difference in 90-day survival between each subgroup (ACDHF: 41.7%, de novo: 83.3%, *P* = .057).

**Table 4.**
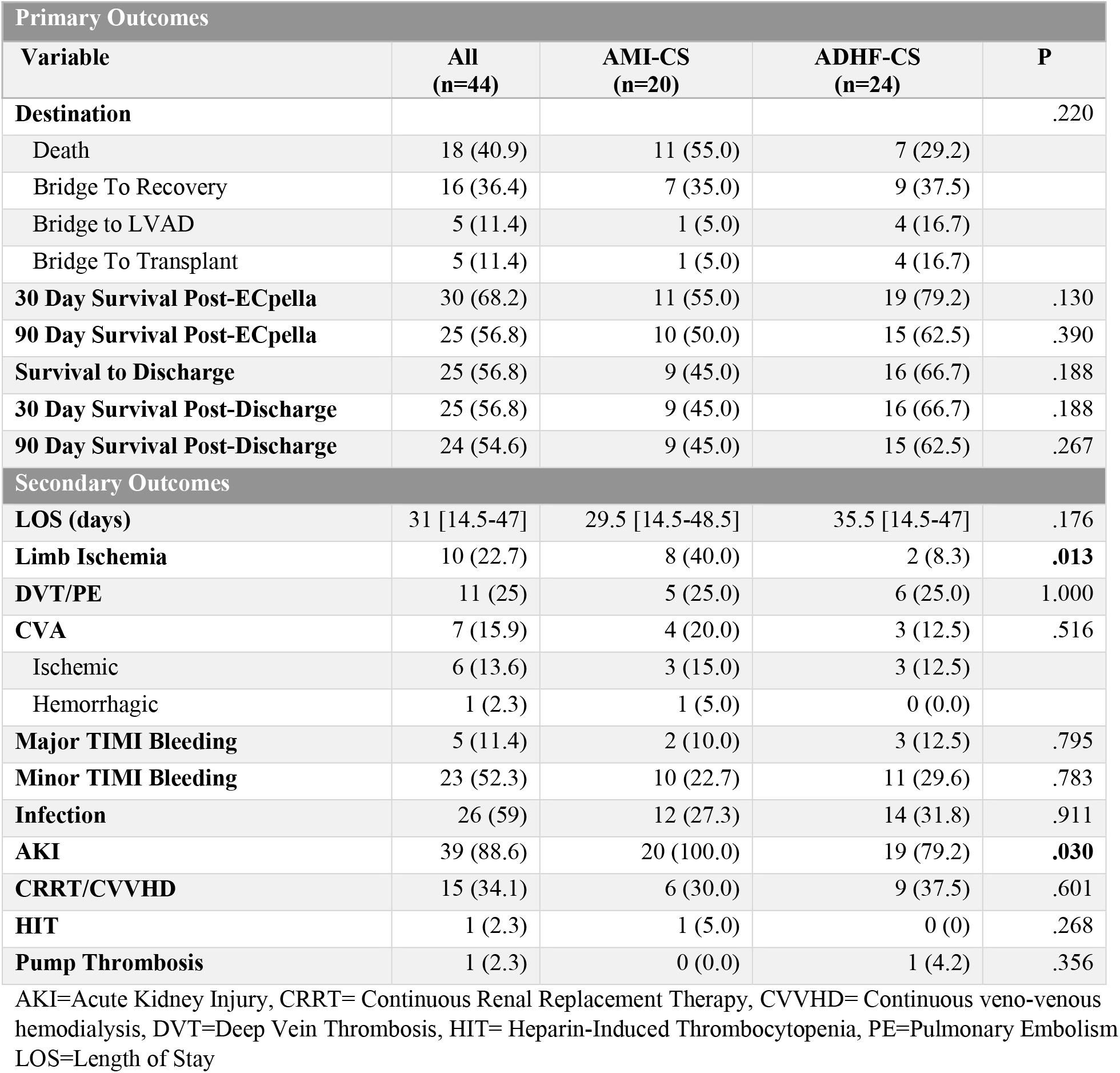
Primary and Secondary Outcomes by CS Etiology.

**Table 5.**
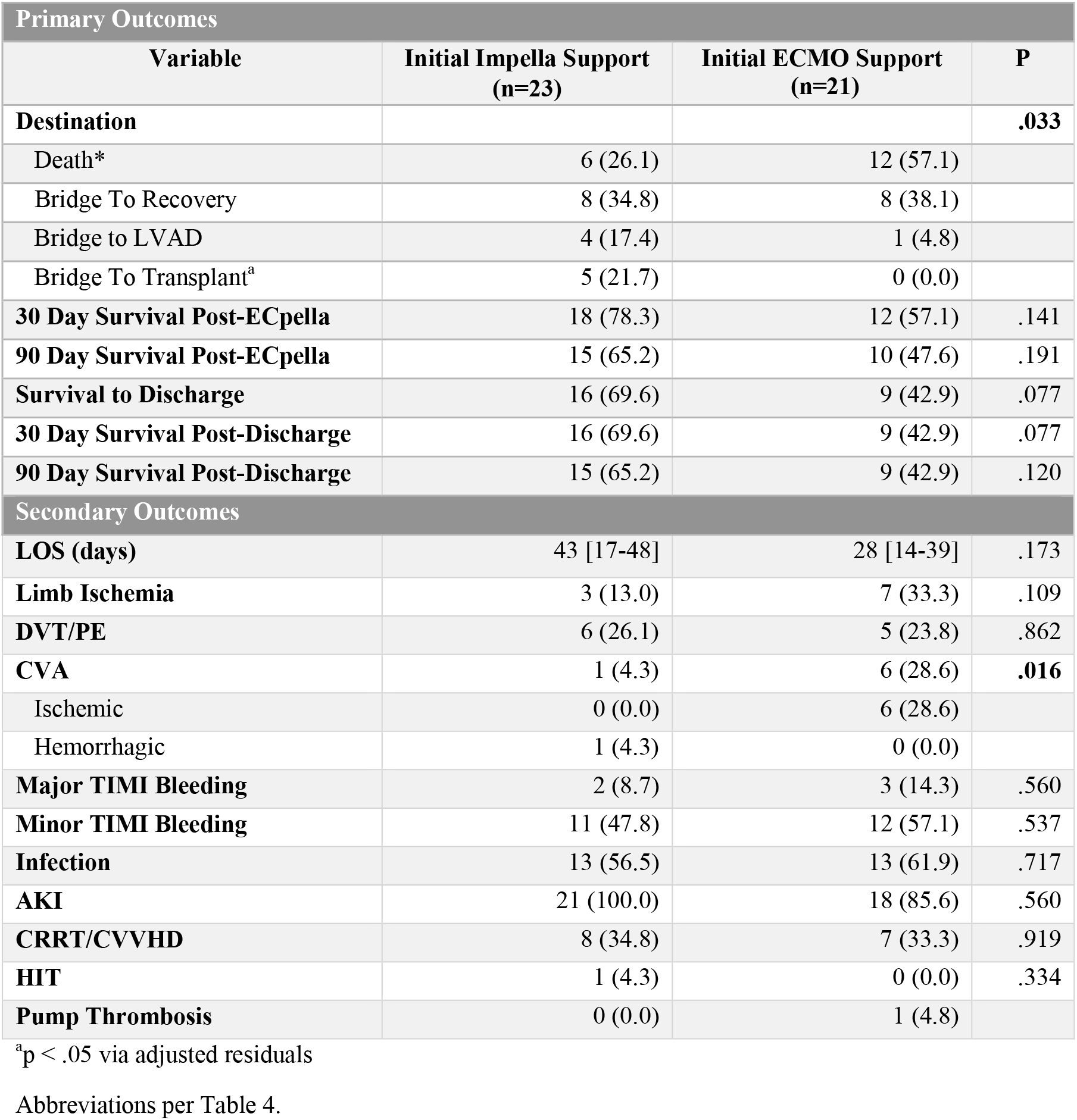
Primary and Secondary Outcomes by Initial Support Strategy.

| Primary Outcomes |  |  |  |
| --- | --- | --- | --- |
| Variable | Initial Impella Support<br>(n=23) | Initial ECMO Support<br>(n=21) | P |
| **Destination** |  |  | **.033** |
| Death* | 6 (26.1) | 12 (57.1) |  |
| Bridge To Recovery | 8 (34.8) | 8 (38.1) |  |
| Bridge to LVAD | 4 (17.4) | 1 (4.8) |  |
| Bridge To Transplant<sup>a</sup> | 5 (21.7) | 0 (0.0) |  |
| **30 Day Survival Post-ECpella** | 18 (78.3) | 12 (57.1) | .141 |
| **90 Day Survival Post-ECpella** | 15 (65.2) | 10 (47.6) | .191 |
| **Survival to Discharge** | 16 (69.6) | 9 (42.9) | .077 |
| **30 Day Survival Post-Discharge** | 16 (69.6) | 9 (42.9) | .077 |
| **90 Day Survival Post-Discharge** | 15 (65.2) | 9 (42.9) | .120 |
| Secondary Outcomes |  |  |  |
| **LOS (days)** | 43 [17-48] | 28 [14-39] | .173 |
| **Limb Ischemia** | 3 (13.0) | 7 (33.3) | .109 |
| **DVT/PE** | 6 (26.1) | 5 (23.8) | .862 |
| **CVA** | 1 (4.3) | 6 (28.6) | **.016** |
| Ischemic | 0 (0.0) | 6 (28.6) |  |
| Hemorrhagic | 1 (4.3) | 0 (0.0) |  |
| **Major TIMI Bleeding** | 2 (8.7) | 3 (14.3) | .560 |
| **Minor TIMI Bleeding** | 11 (47.8) | 12 (57.1) | .537 |
| **Infection** | 13 (56.5) | 13 (61.9) | .717 |
| **AKI** | 21 (100.0) | 18 (85.6) | .560 |
| **CRRT/CVVHD** | 8 (34.8) | 7 (33.3) | .919 |
| **HIT** | 1 (4.3) | 0 (0.0) | .334 |
| **Pump Thrombosis** | 0 (0.0) | 1 (4.8) |  |

<sup>a</sup>p < .05 via adjusted residuals
Abbreviations per Table 4.

**Table 6.**
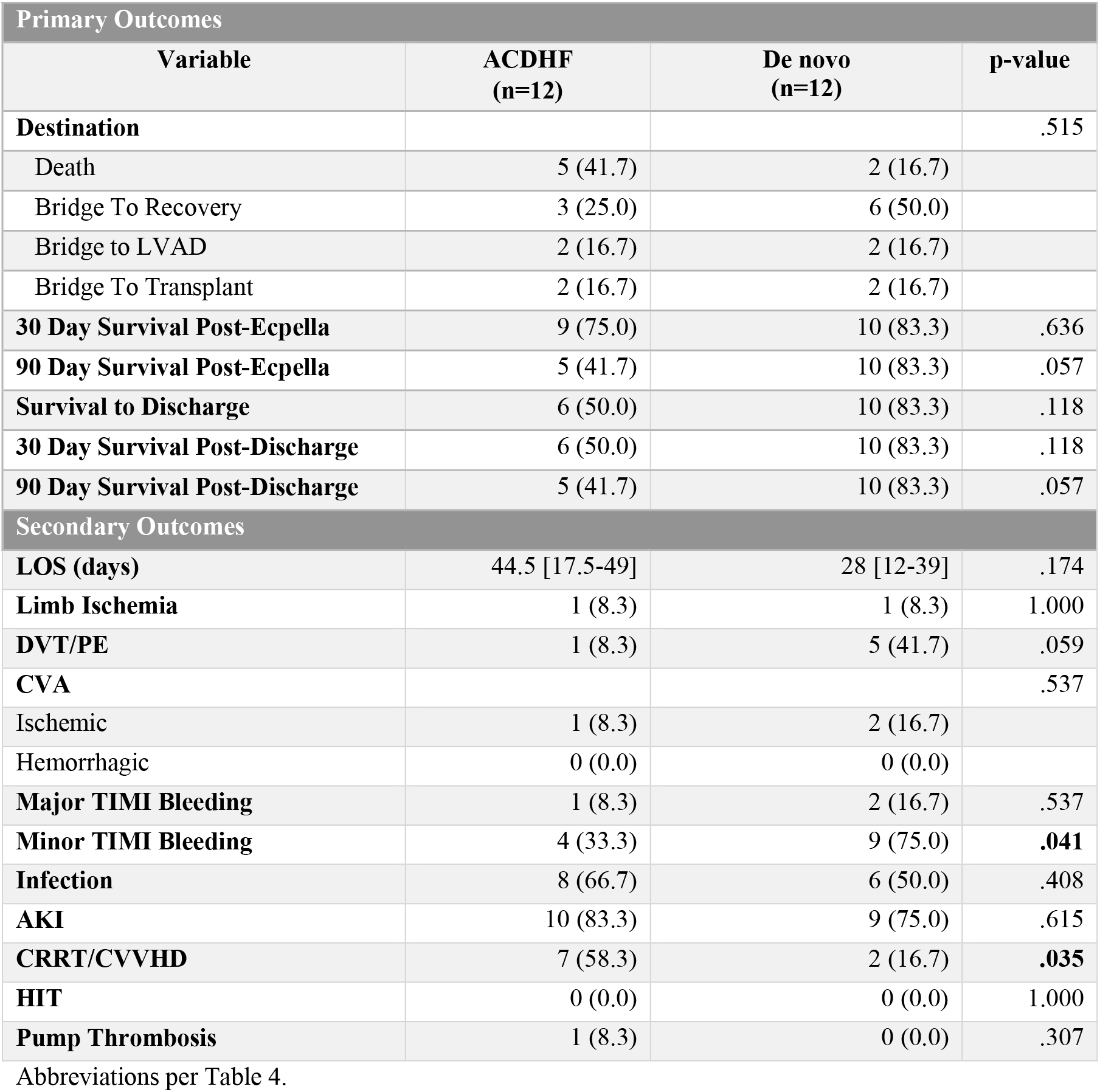
Primary and Secondary Outcomes: ACDHF vs. de novo HF.

| Primary Outcomes |  |  |  |
| --- | --- | --- | --- |
| Variable | ACDHF<br>(n=12) | De novo<br>(n=12) | p-value |
| **Destination** |  |  | .515 |
| Death | 5 (41.7) | 2 (16.7) |  |
| Bridge To Recovery | 3 (25.0) | 6 (50.0) |  |
| Bridge to LVAD | 2 (16.7) | 2 (16.7) |  |
| Bridge To Transplant | 2 (16.7) | 2 (16.7) |  |
| **30 Day Survival Post-Ecpella** | 9 (75.0) | 10 (83.3) | .636 |
| **90 Day Survival Post-Ecpella** | 5 (41.7) | 10 (83.3) | .057 |
| **Survival to Discharge** | 6 (50.0) | 10 (83.3) | .118 |
| **30 Day Survival Post-Discharge** | 6 (50.0) | 10 (83.3) | .118 |
| **90 Day Survival Post-Discharge** | 5 (41.7) | 10 (83.3) | .057 |
| Secondary Outcomes |  |  |  |
| **LOS (days)** | 44.5 [17.5-49] | 28 [12-39] | .174 |
| **Limb Ischemia** | 1 (8.3) | 1 (8.3) | 1.000 |
| **DVT/PE** | 1 (8.3) | 5 (41.7) | .059 |
| **CVA** |  |  | .537 |
| Ischemic | 1 (8.3) | 2 (16.7) |  |
| Hemorrhagic | 0 (0.0) | 0 (0.0) |  |
| **Major TIMI Bleeding** | 1 (8.3) | 2 (16.7) | .537 |
| **Minor TIMI Bleeding** | 4 (33.3) | 9 (75.0) | .041 |
| **Infection** | 8 (66.7) | 6 (50.0) | .408 |
| **AKI** | 10 (83.3) | 9 (75.0) | .615 |
| **CRRT/CVVHD** | 7 (58.3) | 2 (16.7) | .035 |
| **HIT** | 0 (0.0) | 0 (0.0) | 1.000 |
| **Pump Thrombosis** | 1 (8.3) | 0 (0.0) | .307 |

Abbreviations per Table 4.

In our study population, 36.4% of patients recovered after ECpella support without additional intervention while 11.4% of patients were bridged to LVAD support and another 11.4% of patients were bridged to heart transplant. Although patients with ADHF-CS had a higher proportion of patients that were bridged to LVAD and heart transplant, there was no statistically significant difference in destination between the AMI-CS and ADHF-CS (*P* = .220). Patients that received Impella support initially were more likely to bridge to transplant (Impella: 21.7%, VA-ECMO: 0.0%, *P* < .05) whereas patients with initial VA-ECMO support were more likely to experience death on ECpella support (Impella: 26.1%, VA-ECMO: 57.1%, *P <* .05). Among patients with ADHF-CS, there was no significant difference in destination between both ACDHF and de novo heart failure groups, although there was a larger proportion of de novo heart failure patients that were bridged to recovery (ACDHF: 25.0%, de novo: 50%, *P* = .515).

The overall median LOS for all patients was 31 days with no significant difference between the AMI-CS cohort and the ADHF-CS cohort (AMI-CS: 29.5 [14.5-48.5], ADHF-CS: 35.5 [14.5-47], *P* = .176) and no difference in LOS by initial support strategy (Impella: 43 [17-48], VA-ECMO: 28 [14-39], *P* = .173). Patients with AMI-CS were more likely to sustain limb ischemia (AMI-CS: 40%; ADHF-CS: 8.3%, *P* = .013) and acute kidney injury (AMI-CS: 100.0%; ADHF-CS: 79.2%, *P* = .030) in comparison to ADHF-CS patients. Patients supported initially by VA-ECMO had a higher rate of ischemic CVA (Impella: 0%, VA-ECMO: 28.6%, *P=.*016) when compared to patients supported initially with Impella. When comparing patients with ACDHF and de novo heart failure, patients with ACDHF had a higher proportion of patients requiring CRRT (ACDHF: 58.3%, de novo: 16.7%, *P* = .035) while the de novo heart failure patient population had a higher proportion of patients with minor TIMI bleeding (ACDHF: 33.3%, de novo: 75.0%, *P* = .041).

## Discussion

To our knowledge, this study is one of the largest single-center studies of ECpella support in CS and the only study that characterizes ECpella utilization across CS etiology and by initial support strategy. The overall 90-day post-discharge survival of all CS patients was 54.6%, which illustrates the vulnerability of the CS population despite maximal cardiac support. ECpella support was employed in a severely ill cohort of cardiogenic shock patients in this study, with patients with AMI-CS presenting with a median lactate of 9.1. There was no difference in 90-day survival or destination after support between the AMI-CS and ADHF-CS cohorts and stratification of the ADHF-CS cohort into ACDHF and de novo heart failure groups did not reveal a significant difference in survival or destination. However, patients with an initial Impella support strategy were more likely to survive ECpella support and be bridged to a heart transplant when compared to patients with initial VA-ECMO support.

To date, there is only a limited number of retrospective analyses studying the utilization of ECpella support in CS, which highlights its novelty as a support strategy. Similar survival rates between 43%-53% with early ECpella support have been noted in retrospective single center studies comparing VA-ECMO and IABP support to ECpella support for AMI-CS and cardiac arrest^14,15^ and in both single center and multicenter retrospective analyses comparing VA-ECMO support to ECpella support for both AMI-CS and ADHF-CS patients^10,11^. In these studies, the initiation of ECpella support was performed on patients with existing VA ECMO support to assist in left ventricular unloading and prevent left ventricular dilatation and thrombus formation^8^. Our study is the first to also include patients with initial Impella support that received subsequent VA-ECMO support for cardiac output augmentation.

AMI-CS and ADHF-CS are two distinct phenotypes of CS with differing pathophysiology^16^. AMI-CS patients undergo an abrupt reduction in functional myocardium requiring an extensive period of ventricular remodeling and are heavily dependent on revascularization for rescue of viable myocardium^17,18^. The pathophysiology of ADHF-CS is incompletely understood and has been attributed to multiple factors including prior comorbidities, prior myocardial damage from previous ischemic insults or inflammatory processes such as myocarditis, and neurohormonal activation states affecting arterial and venous vascular tone^19^. Limited evidence exists regarding the differing mortality between these two cohorts, however a recent single-center study by Sinha et al demonstrated that ADHF-CS patients have a lower 1-year mortality compared to AMI-CS patients^16^. This difference was attributed to ADHF-CS patients possessing a higher tolerance of lower cardiac output states due to development of chronic compensation, although this is only applicable to patients with ACDHF rather than patients presenting with de novo heart failure ^16,20^.

The lack of significant difference in survival and destination to advanced therapies in our study between the ADHF-CS cohort and AMI-CS cohort is multifactorial. Despite their differing pathophysiology, approximately 80% of the patients in this study presented with SCAI Stage D/E shock, suggesting that end-organ damage had already occurred prior to full cardiac support with ECpella. This is supported by almost 90% of allcomers in cardiogenic shock presenting with acute kidney injury prior to ECpella initiation. These findings contrast with the prior study by Sinha et al in which only 10% of allcomers with cardiogenic shock received ECpella support and 50% of patients presented with SCAI Stage C shock. Although there was a trend towards increased survival in the ADHF-CS cohort at 90 days post ECpella initiation (ADHF-CS: 62.5%, AMI-CS: 50%, NNT:8.0), durable LVAD implantation (ADHF-CS: 16.7%, AMI-CS: 5%, NNT: 8.5), and heart transplant (ADHF-CS: 16.7%, AMI-CS: 5%, NNT: 8.5), the lack of significant differences may be attributed to the study being underpowered to detect true differences, if they exist (**Fig. 1**, **Fig. 2)**. As the use of ECpella support continues to increase for refractory cardiogenic shock, further research with larger sample sizes is needed to delineate survival and destination differences between these two cohorts.

**Figure 1.**
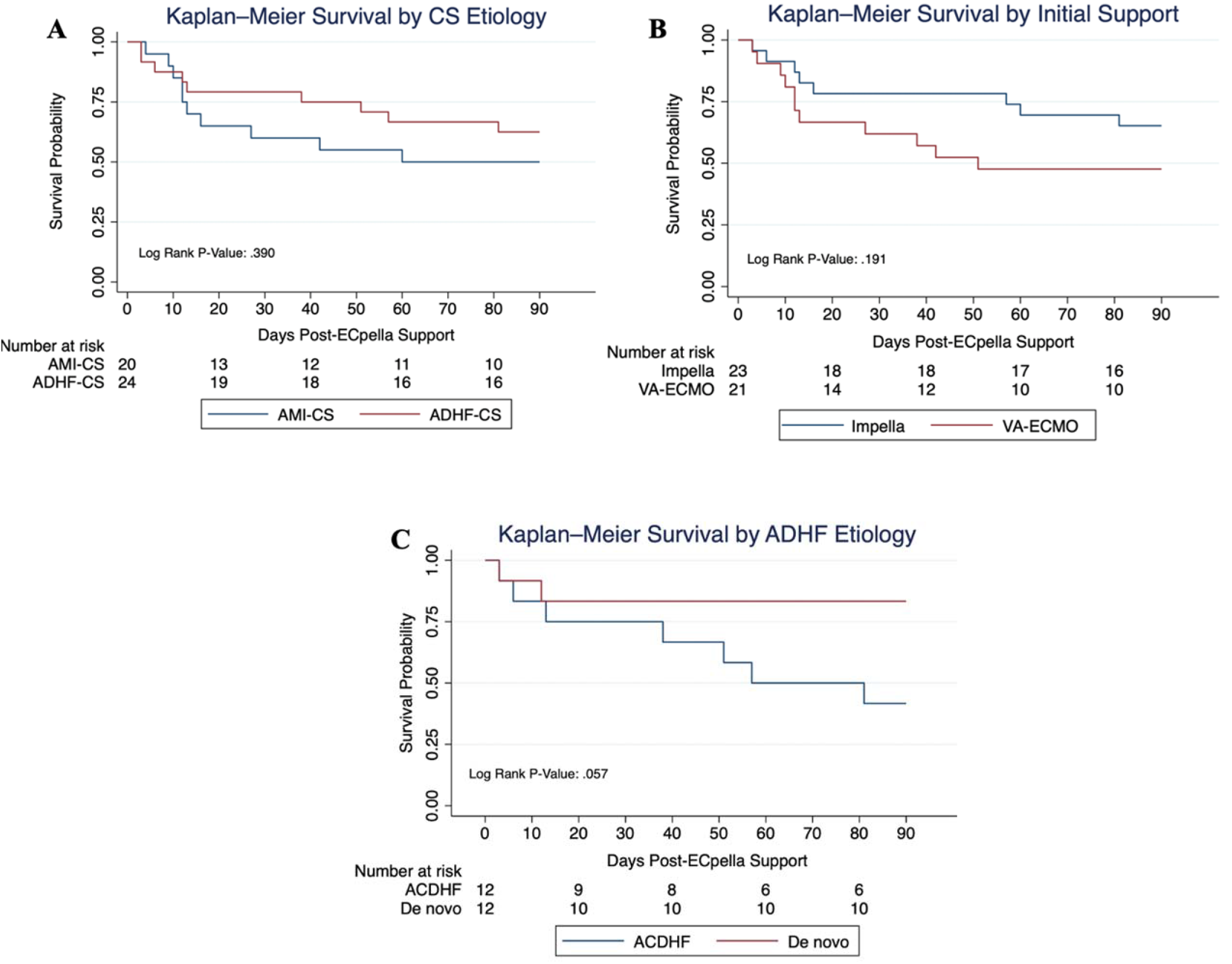
CS Etiology and Initial Support do not affect 90-day survival. A) 62.5% (n=16) of patients with ADHF-CS survived after 90-days post ECpella support compared to 45% (n=9) of patients with AMI-CS. B) 62.5% (n=15) of patients with initial Impella support survived after 90-days post ECpella support compared to 47.6% (n=10) of patients with initial VA-ECMO support. C) ADHF-CS sub-cohorts, in which 83.3% (n=10) of patients with de novo heart failure survived after 90-days post-ECpella support compared to 41.7% (n=5) of patients with acute-on-chronic decompensated heart failure.

**Figure 2.**
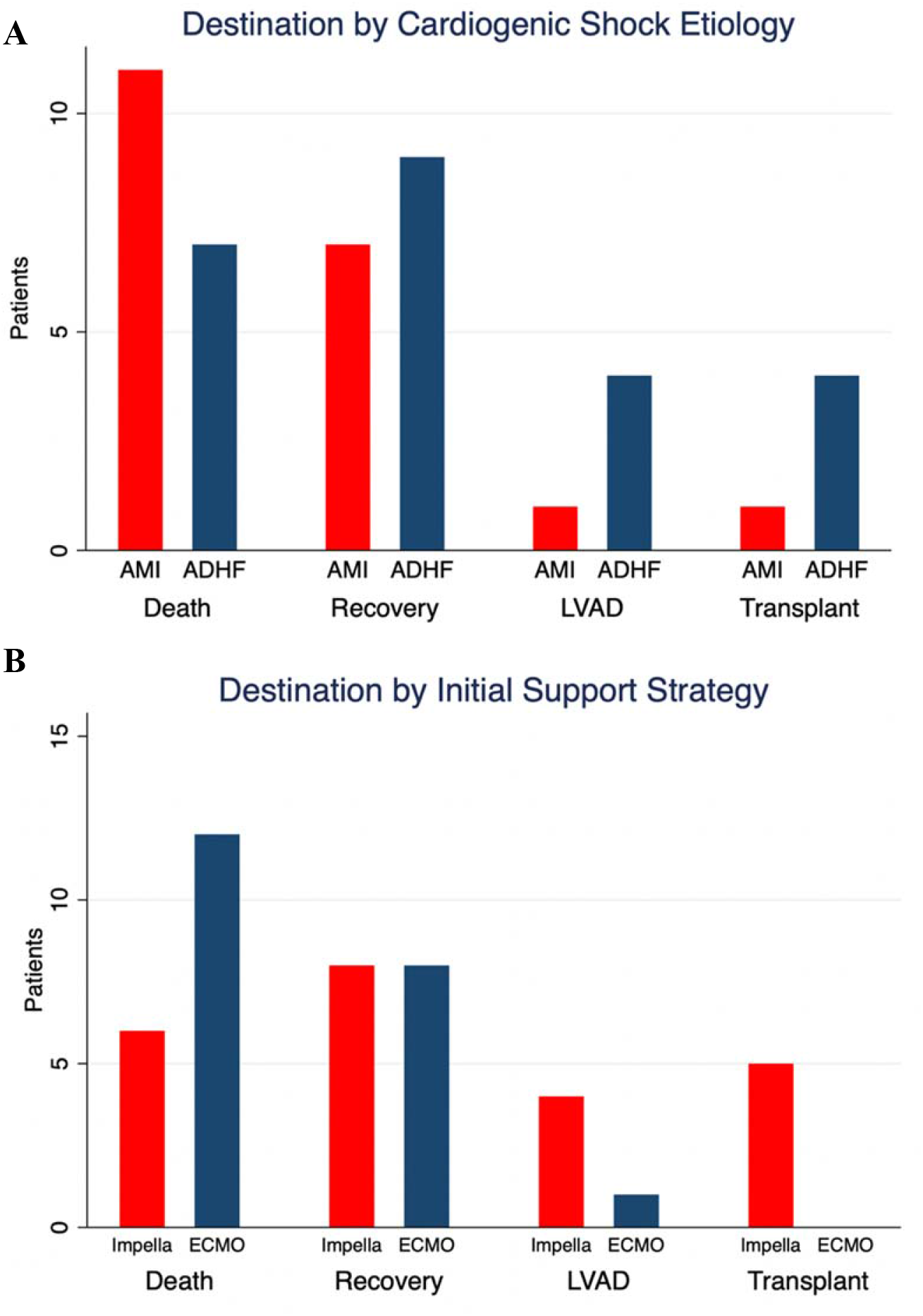
Destination does not vary with CS Etiology but varies with initial support. A) Despite a higher proportion of AMI-CS patients that died while on ECpella support (55%, n=11) compared to ADHF-CS patients (29%, n=7), there was no statistical significance in destination between the two cohorts (*p* =.220). B) Patients on initial VA-ECMO support had a significantly higher proportion that died (n=12, 57.1%) while on ECpella support compared to initial Impella support (n=6, 26.1%). Patients initially supported on Impella were more likely to be bridged to transplant.

Regarding secondary outcomes, patients with AMI-CS were more likely to sustain limb ischemia requiring surgical intervention and all AMI-CS patients sustained acute kidney injury during their hospital admission. These outcomes are likely secondary to lack of compensatory mechanisms to lower cardiac output states that have been demonstrated in patients with ADHF-CS due to long term adaptation to LV dilation and higher filling pressures^16,21^. In addition, despite 0% of AMI-CS patients having a prior diagnosis of PAD, we suspect this is likely due to underdiagnosis and these patients likely had higher burden of atherosclerosis leading to a higher incidence of limb ischemia in this cohort. Furthermore, 10% of patients with AMI-CS were unable to get coronary intervention due to significant shock despite ECpella support, which likely exacerbated these complications.

Patients with initial Impella support were more likely to survive ECpella support and be bridged to transplant in comparison to patients with initial VA ECMO support despite having similar SCAI stages on presentation (**Fig. 1**, **Fig. 2**). A recent meta-analysis by Batchelor et al demonstrated that patients with AMI-CS (n=7093) that were treated with Impella support rather than VA-ECMO support had reduced short- and medium-term mortality which was attributed to the initial increased afterload with VA-ECMO limiting myocardial recovery^22^. Though the median for Impella placement after VA ECMO support in this study was one day, recent studies have shown that left ventricular venting via Impella prior to 12 hours of support is associated with reduced short-term mortality^23,24^. Therefore, this initial increase in afterload to the left ventricle may have limited initial myocardial recovery and resulted in additional complications that affected mortality and eventual destination.

Patients initially supported with VA ECMO had statistically significant anemia and thrombocytopenia prior to full ECpella support which could be secondary to blood loss during cannulation or hemolysis from ECMO support. This relative anemia and thrombocytopenia could have led to a component of superimposed hemorrhagic shock in these patients and potentially decreased oxygen delivery. In addition, ischemic stroke is known to be a risk with patients on VA ECMO^25,26^ and patients with initial VA ECMO support had a significant higher rate of ischemic stroke compared to patients with initial Impella support. Aside from the sequalae after an ischemic stroke, patients who sustained ischemic stroke likely did not qualify for advanced therapies which likely led to a higher mortality in the initial VA ECMO cohort. A higher proportion of initial VA ECMO patients received Impella 5.5 support rather than Impella CP support which has a peak flow rate of 6 L/min compared to 4.3 L/min respectively. Though this trend was likely due to ease of axillary surgical placement of Impella 5.5 in patients cannulated with VA ECMO, this augmented support did not improve survival.

In the sub-analysis of ADHF-CS patients, patients with de novo heart failure did not have differing survival or destination to advanced therapies when compared to patients with ACDHF patients despite being younger and having less comorbidities. Though patients with de novo heart failure trended towards increased survival at 90 days post discharge (de novo: 83.3%, ACDHF: 41.7%; NNT: 2.4), the lack of significant difference may be attributed to the study being underpowered to detect true differences, if they exist (**Fig. 1**). Patients with ACDHF have been hypothesized to have physiologic adaptations that preserve stroke volume with lower LVEF and have even been shown to have different myosin structure in diaphragmatic muscles to assist oxidative capacity ^21^. These compensatory mechanisms have led to ACDHF patients having similar and even lower short-term mortality rates than those with de novo heart failure in large retrospective studies ^21,27^. Although de novo heart failure patients supported on ECpella trended towards higher survival at 90 days post discharge, further studies with larger sample sizes in these groups are required to minimize the potential of Type II error in this cohort.

Patients with ACDHF were more likely to require CVVHD or CRRT compared to patients with de novo heart failure, while patients with de novo heart failure had higher minor bleeding events as classified by TIMI. The greater progression to CVVHD/CRRT in the ACDHF group was likely secondary to ACDHF having a higher proportion of patients with CKD prior to ECpella support. Given that TIMI criteria for minor bleeding is based off hemoglobin of ≥ 3 g/dL, it is unclear if patients with de novo heart failure had higher bleeding events or initially had hemodilution from greater volume resuscitation due to initial undifferentiated shock, which warrants further investigation.

### Limitations

Despite this study’s strengths as one of largest single center studies evaluating ECpella across AMI-CS and ADHF-CS as well as the first study to describe patients with initial Impella support that received subsequent VA-ECMO support for cardiogenic shock, there are several limitations that should be acknowledged. To begin, given that the use of ECpella support is often limited to a subset of cardiogenic shock patients that are extremely ill, the low sample size of each cohort may have contributed to the study being underpowered to detect true differences between each group. The duration of chronic heart failure prior to presentation is unknown in the ACDHF cohort, and varying chronicity of heart failure is a factor not accounted for in survival analysis of this study. Our study also had a lack of diversity with a higher proportion of white males, which affects the generalizability of our results to the greater cardiogenic shock population. Lastly, given that the study is retrospective, non-randomized and only involves a single center, the study is subject to potential selection bias and subject to center-specific practice patterns in cardiogenic shock management. Future multicenter, randomized, adequately powered, prospective studies are necessary to validate our results and analyze all associations related to ECpella support for different etiologies of cardiogenic shock.

## Conclusion

ECpella support is a feasible support strategy for both AMI-CS and ADHF-CS with no difference in survival rates or destination to advance therapies. AMI-CS patients are more prone to sustaining acute kidney injury and limb ischemia requiring surgical intervention as complications of support. When comparing initial support strategy for ECpella, patients with initial Impella support are more likely to survive ECpella support and bridge to transplant, while also having a lower rate of acute ischemic stroke. No survival or destination differences are noted between ACDHF patients and de novo heart failure patients although patients with ACDHF are more likely to have a history of CKD and require CRRT/CVVHD while on ECpella support. Future larger multicenter studies are required to better discern the differences between AMI-CS and ADHF-CS on ECpella support.

## Data Availability

Patients were identified through internal review of an internal ECMO and Impella database maintained by perfusion services at University of Pittsburgh Presbyterian Hospital between December 2020 to January 2023. Data is not shared openly to protect patient confidentiality.

## Non-standard Abbreviations and Acronyms

ACDHF: Acute-on-chronic decompensated heart failure
ADHF-CS: Acute decompensated heart failure complicated by cardiogenic shock
AMI-CS: Acute myocardial infarction complicated by cardiogenic shock
CS: Cardiogenic shock
DCI: Distressed Communities’ Index
ECpella: VA-ECMO and concomitant Impella support
VA-ECMO: Veno-arterial Extracorporeal Membrane Oxygenation

## References

1. van Diepen S, Katz JN, Albert NM, Henry TD, Jacobs AK, Kapur NK, Kilic A, Menon V, Ohman EM, Sweitzer NK, Thiele H, Washam JB, Cohen MG. Contemporary Management of Cardiogenic Shock: A Scientific Statement From the American Heart Association. Circulation. 2017;136:e232–e268.

2. Wayangankar SA, Bangalore S, McCoy LA, Jneid H, Latif F, Karrowni W, Charitakis K, Feldman DN, Dakik HA, Mauri L, Peterson ED, Messenger J, Roe M, Mukherjee D, Klein A. Temporal Trends and Outcomes of Patients Undergoing Percutaneous Coronary Interventions for Cardiogenic Shock in the Setting of Acute Myocardial Infarction: A Report From the CathPCI Registry. JACC Cardiovasc Interv. 2016;9:341–351.

3. Adams KF, Fonarow GC, Emerman CL, LeJemtel TH, Costanzo MR, Abraham WT, Berkowitz RL, Galvao M, Horton DP, ADHERE Scientific Advisory Committee and Investigators. Characteristics and outcomes of patients hospitalized for heart failure in the United States: rationale, design, and preliminary observations from the first 100,000 cases in the Acute Decompensated Heart Failure National Registry (ADHERE). Am Heart J. 2005;149:209–216.

4. Shah M, Patnaik S, Patel B, Ram P, Garg L, Agarwal M, Agrawal S, Arora S, Patel N, Wald J, Jorde UP. Trends in mechanical circulatory support use and hospital mortality among patients with acute myocardial infarction and non-infarction related cardiogenic shock in the United States. Clin Res Cardiol. 2018;107:287–303.

5. Vallabhajosyula S, Prasad A, Bell MR, Sandhu GS, Eleid MF, Dunlay SM, Schears GJ, Stulak JM, Singh M, Gersh BJ, Jaffe AS, Holmes DR, Rihal CS, Barsness GW. Extracorporeal Membrane Oxygenation Use in Acute Myocardial Infarction in the United States, 2000 to 2014. Circ Heart Fail. 2019;12:e005929.

6. Dickstein ML. The Starling Relationship and Veno-Arterial ECMO: Ventricular Distension Explained. ASAIO J. 2018;64:497–501.

7. Donker DW, Brodie D, Henriques JPS, Broomé M. Left ventricular unloading during veno-arterial ECMO: a review of percutaneous and surgical unloading interventions. Perfusion. 2019;34:98–105.

8. Belohlavek J, Hunziker P, Donker DW. Left ventricular unloading and the role of ECpella. European Heart Journal Supplements. 2021;23:A27–A34.

9. Truby LK, Takeda K, Mauro C, Yuzefpolskaya M, Garan AR, Kirtane AJ, Topkara VK, Abrams D, Brodie D, Colombo PC, Naka Y, Takayama H. Incidence and Implications of Left Ventricular Distention During Venoarterial Extracorporeal Membrane Oxygenation Support. ASAIO J. 2017;63:257–265.

10. Pappalardo F, Schulte C, Pieri M, Schrage B, Contri R, Soeffker G, Greco T, Lembo R, Müllerleile K, Colombo A, Sydow K, De Bonis M, Wagner F, Reichenspurner H, Blankenberg S, Zangrillo A, Westermann D. Concomitant implantation of Impella® on top of veno-arterial extracorporeal membrane oxygenation may improve survival of patients with cardiogenic shock. European Journal of Heart Failure. 2017;19:404–412.

11. Patel SM, Lipinski J, Al-Kindi SG, Patel T, Saric P, Li J, Nadeem F, Ladas T, Alaiti A, Phillips A, Medalion B, Deo S, Elgudin Y, Costa MA, Osman MN, Attizzani GF, Oliveira GH, Sareyyupoglu B, Bezerra HG. Simultaneous Venoarterial Extracorporeal Membrane Oxygenation and Percutaneous Left Ventricular Decompression Therapy with Impella Is Associated with Improved Outcomes in Refractory Cardiogenic Shock. ASAIO Journal. 2019;65:21.

12. Grajeda Silvestri ER, Pino JE, Donath E, Torres P, Chait R, Ghumman W. Impella to unload the left ventricle in patients undergoing venoarterial extracorporeal membrane oxygenation for cardiogenic shock: A systematic review and meta-analysis. J Card Surg. 2020;35:1237– 1242.

13. Baran DA, Grines CL, Bailey S, Burkhoff D, Hall SA, Henry TD, Hollenberg SM, Kapur NK, O’Neill W, Ornato JP, Stelling K, Thiele H, van Diepen S, Naidu SS. SCAI clinical expert consensus statement on the classification of cardiogenic shock: This document was endorsed by the American College of Cardiology (ACC), the American Heart Association (AHA), the Society of Critical Care Medicine (SCCM), and the Society of Thoracic Surgeons (STS) in April 2019. Catheter Cardiovasc Interv. 2019;94:29–37.

14. Shibasaki I, Masawa T, Abe S, Ogawa H, Takei Y, Tezuka M, Seki M, Kato T, Watanabe R, Koshiji N, Saitou S, Ogata K, Haruyama Y, Toyoda S, Fukuda H. Benefit of veno-arterial extracorporeal membrane oxygenation combined with Impella (ECpella) therapy in acute coronary syndrome with cardiogenic shock. Journal of Cardiology. 2022;80:116–124.

15. Unoki T, Kamentani M, Nakayama T, Tamura Y, Konami Y, Suzuyama H, Inoue M, Yamamuro M, Taguchi E, Sawamura T, Nakao K, Sakamoto T. Impact of extracorporeal CPR with transcatheter heart pump support (ECPELLA) on improvement of short-term survival and neurological outcome in patients with refractory cardiac arrest – A single-site retrospective cohort study. Resuscitation Plus. 2022;10:100244.

16. Sinha SS, Rosner CM, Tehrani BN, Maini A, Truesdell AG, Lee SB, Bagchi P, Cameron J, Damluji AA, Desai M, Desai SS, Epps KC, deFilippi C, Flanagan MC, Genovese L, Moukhachen H, Park JJ, Psotka MA, Raja A, Shah P, Sherwood MW, Singh R, Tang D, Young KD, Welch T, O’Connor CM, Batchelor WB. Cardiogenic Shock From Heart Failure Versus Acute Myocardial Infarction: Clinical Characteristics, Hospital Course, and 1-Year Outcomes. Circulation: Heart Failure. 2022;15:e009279.

17. Hochman JS, Sleeper LA, Webb JG, Dzavik V, Buller CE, Aylward P, Col J, White HD. Early Revascularization Improves Long-Term Survival for Cardiogenic Shock Complicating Acute Myocardial Infarction. JAMA. 2006;295:2511–2515.

18. Garza MA, Wason EA, Zhang JQ. Cardiac remodeling and physical training post myocardial infarction. World J Cardiol. 2015;7:52–64.

19. Njoroge JN, Teerlink JR. Pathophysiology and Therapeutic Approaches to Acute Decompensated Heart Failure. Circulation Research. 2021;128:1468–1486.

20. Hummel A, Empen K, Dörr M, Felix SB. De Novo Acute Heart Failure and Acutely Decompensated Chronic Heart Failure. Dtsch Arztebl Int. 2015;112:298–310.

21. Bhatt AS, Berg DD, Bohula EA, Alviar CL, Baird-Zars VM, Barnett CF, Burke JA, Carnicelli AP, Chaudhry S-P, Daniels LB, Fang JC, Fordyce CB, Gerber DA, Guo J, Jentzer JC, Katz JN, Keller N, Kontos MC, Lawler PR, Menon V, Metkus TS, Nativi-Nicolau J, Phreaner N, Roswell RO, Sinha SS, Jeffrey Snell R, Solomon MA, Van Diepen S, Morrow DA. De Novo vs Acute-on-Chronic Presentations of Heart Failure-Related Cardiogenic Shock: Insights from the Critical Care Cardiology Trials Network Registry. Journal of Cardiac Failure. 2021;27:1073–1081.

22. Batchelor RJ, Wheelahan A, Zheng WC, Stub D, Yang Y, Chan W. Impella versus Venoarterial Extracorporeal Membrane Oxygenation for Acute Myocardial Infarction Cardiogenic Shock: A Systematic Review and Meta-Analysis. Journal of Clinical Medicine. 2022;11:3955.

23. Upadhrasta S, Museedi A, Thannoun T, Chaanine AH, Le Jemtel TH. Early Mechanical Circulatory Support for Cardiogenic Shock. Cardiology in Review.;:10.1097/CRD.0000000000000485.

24. Al-Fares AA, Randhawa VK, Englesakis M, McDonald MA, Nagpal AD, Estep JD, Soltesz EG, Fan E. Optimal Strategy and Timing of Left Ventricular Venting During Veno-Arterial Extracorporeal Life Support for Adults in Cardiogenic Shock: A Systematic Review and Meta-Analysis. Circ Heart Fail. 2019;12:e006486.

25. Cappannoli L, Galli M, Zito A, Restivo A, Princi G, Laborante R, Vergallo R, Romagnoli E, Leone AM, Aurigemma C, Massetti M, Sanna T, Trani C, Burzotta F, Savarese G, Crea F, D’Amario D. Venoarterial extracorporeal membrane oxygenation (VA-ECMO) with vs. without left ventricular unloading by Impella: a systematic review and meta-analysis. European Heart Journal - Quality of Care and Clinical Outcomes. 2022;:qcac076.

26. Le Guennec L, Cholet C, Huang F, Schmidt M, Bréchot N, Hékimian G, Besset S, Lebreton G, Nieszkowska A, Leprince P, Combes A, Luyt C-E. Ischemic and hemorrhagic brain injury during venoarterial-extracorporeal membrane oxygenation. Ann Intensive Care. 2018;8:129.

27. Younis A, Mulla W, Goldkorn R, Klempfner R, Peled Y, Arad M, Freimark D, Goldenberg I. Differences in Mortality of New-Onset (De-Novo) Acute Heart Failure Versus Acute Decompensated Chronic Heart Failure. The American Journal of Cardiology. 2019;124:554– 559.

